# Cost-effectiveness analysis of BNT162b2 COVID-19 booster vaccination in the United States

**DOI:** 10.1101/2021.11.14.21266318

**Authors:** Rui Li, Hanting Liu, Christopher K Fairley, Zhuoru Zou, Li Xie, Xinghui Li, Mingwang Shen, Yan Li, Lei Zhang

## Abstract

**Background:** Over 86% of older adults aged ≥65 years are fully vaccinated against SARS-COV-2 in the United States (US). Waning protection of the existing vaccines promotes the new vaccination strategies, such as providing a booster shot for those fully vaccinated.

**Methods:** We developed a decision-analytic Markov model of COVID-19 to evaluate the cost-effectiveness of a booster strategy of Pfizer-BioNTech BNT162b2 (administered 6 months after 2^nd^ dose) in those aged ≥65 years, from a healthcare system perspective.

**Findings:** Compared with 2-doses of BNT162b2 without a booster, the booster strategy in a 100,000 cohort of older adults would incur an additional cost of $3.4 million, but save $6.7 million in direct medical costs in 180 days. This corresponds to a benefit-cost ratio of 1.95 and a net monetary benefit of $3.4 million. Probabilistic sensitivity analysis indicates that with a COVID-19 incidence of 9.1/100,000 person-day, a booster strategy has a high chance (67%) of being cost-effective. The cost-effectiveness of the booster strategy is highly sensitive to the population incidence of COVID-19, with a cost-effectiveness threshold of 8.1/100,000 person-day. This threshold will increase with a decrease in vaccine and booster efficacies. Doubling the vaccination cost or halving the medical cost for COVID-19 treatment alone would not alter the conclusion of cost-effectiveness, but certain combinations of the two might render the booster strategy not cost-effective.

**Interpretation:** Offering BNT162b2 boosters to older adults aged ≥65 years in the US is likely to be cost-effective. Less efficacious vaccines and boosters may still be cost-effective in settings of high SARS-COV-2 transmission.

**Funding:** National Natural Science Foundation of China. Berlina and Bill Gates Foundation

## Introduction

As of 12^nd^ November 2021, the COVID-19 pandemic has claimed more than 5 million lives around the globe ^1^. The economic costs of the COVID-19 are enormous in many countries ^2-4^. In the United States (US), nearly 47 million people have been infected by COVID-19, resulting in more than 755,000 deaths. At the same time, over 69% of the US population aged ≥12 years has been fully vaccinated, and the percentage has reached 86% among older adults aged ≥65 years ^5^. Although the rapid development and distribution of COVID-19 vaccines brought about hopes of curbing the pandemic ^6,7^, recent reports of a growing number of breakthrough infections have raised serious concerns from the public ^8,9^. Both the rampant transmission of the SARS-COV-2 Delta variant and the waning protection of the existing vaccines likely contribute to the increasing number of breakthrough infections. New vaccination strategies, such as providing vaccine boosters to those who have been fully vaccinated, are currently being discussed ^10^.

The US Food and Drug Administration (FDA) approved the use of a booster shot for those who received the 2-dose Pfizer-BioNTech COVID-19 vaccine on 22nd September 2021. Although the FDA approved the use of booster shots, debate continues as whether booster shots should be offered and, if so, to which populations ^10^. On one hand, real-world data from countries such as Israel showed that booster shots significantly increased the vaccine’s effectiveness, which provides additional protection against COVID-19, especially for older adults and people with compromised immunity ^11^. In addition, recent serological studies have demonstrated a substantial waning of vaccine-elicited immunity against SARS-COV-2 infection 6-11 months after administering its second dose ^12-14^. On the other hand, the wide dissemination of COVID-19 booster shots may further increase the already skyrocketing healthcare costs and exacerbate the health equity issue exposed by the pandemic ^15,16^. There is, thus, an urgent need to analyse the potential impact of widely administering booster shots in the US.

In this study, we aim to, for the first time, assess the cost-effectiveness of the COVID-19 booster strategy in the US. Based on a well-designed decision-analytic Markov model, our findings will aid public health practitioners and policymakers to determine whether universally administering booster shots among those aged ≥65 years who have been fully vaccinated would be a cost-effective strategy. Our study will also explore the key factors that may affect its cost-effectiveness.

## Methods

### Study design

We conducted an economic evaluation on the cost-effectiveness of booster vaccination of Pfizer-BioNTech (6 months after 2^nd^ dose) in those aged ≥65 years, based on a decision-analytic Markov model. The evaluation was conducted from a healthcare system perspective. The model was constructed using TreeAge Pro 2021 R1.1, and the analysis was conducted according to the Consolidated Health Economic Evaluation Reporting Standards statement ^17^.

### Modelling

A decision-analytic Markov model was constructed to simulate the disease progression of SARS-CoV-2 infection in a designated initial cohort of 100,000 individuals aged ≥65 years over a period of 180 days. As the Delta variant of SARS-COV-2 has become dominant in the US, our study mainly focused on modelling the transmission of the Delta variant. Existing evidence indicated that the vaccine efficacy (VE) of Pfizer-BioNTech/ BNT162b2 would gradually wane after six months ^12-14^. Thus, we defined the vaccine efficacy from 2 weeks to 6 months after the 2^nd^ dose of vaccines as a ‘short-term VE’, whereas the vaccine efficacy six months beyond the 2^nd^ dose as a ‘long-term VE’.

The model consisted of 9 health states depicting varied disease progression of COVID-19 (**Figure S1**). A fully vaccinated individual may be infected by SARS-CoV-2 and enter a ‘latent infection state’. After a mean incubation period of 5.2 (4.1-7.0) days ^18^, About 83% of infected individuals developed symptoms ^19^, and the remaining asymptomatic infections would spontaneously recover. A symptomatic infection might first exhibit ‘mild/moderate’ symptoms. It might ‘recover’ or deteriorate to a ‘severe’ state. A patient in the ‘severe’ state might ‘recover’ or progress to the ‘critical’ state. Similarly, a patient in the ‘critical’ state might ‘recover’ or ‘die’. Transition probabilities between states were estimated using the formula *p* = 1 − *e*^−*r*^, where *r* denoted the daily transition rate ^19^. Model cycle length was 1 day, with a half-cycle correction applied. We defined the approach of 2-dose BNT162b2 without booster shot as the ‘baseline’ and evaluated the health and cost benefits of implementing the booster strategy in older adults aged ≥65 years in the US.

### Data collection

We collected information on the vaccine efficacy of BNT162b2 for SARS-CoV-2 (Delta variant) infection in older adults aged ≥65 years based on an ongoing systematic review conducted by The International Vaccine Access Center ^20^. We included 11 eligible studies to estimate the pooled short-term and long-term VE of the 2-dose vaccination and also the VE of the booster shot (Appendix 1.2). Based on the varied VE for preventing COVID-19 infection and severe progression, we developed a mathematical model to estimate the distributions of clinical disease stages after being infected by SARS-CoV-2 in vaccinated individuals, compared to that in unvaccinated ones (Appendix 1.3). We estimated the population incidence of COVID-19 in US older adults to be 9.1/100,000 person-day by averaging out the published data (of US CDC) over the last 180 days before the approval of a booster shot on 22^nd^ September 2021 ^21^ (Appendix 1.4).

The costs of booster vaccination included the cost of BNT162b2 vaccine ($19.5/dose) ^22^ and the vaccination administration ($17.1/dose) ^23,24^. The cost of PCR testing for SARS-CoV-2 infection was estimated to be $51.0/person according to the COVID-19 testing pricing from the medical insurance administrative contractor ^25^. We collected the total direct medical costs from hospitalisation for each COVID-19 clinical stage based on the Projected Economic Impact Report of the US Healthcare System and Health System Tracker ^26,27^. We calculated the corresponding daily cost by dividing the total cost by the duration of the clinical stages (Table S1, Appendix 1.5).

Health utility scores for COVID-19 patients were derived from the disutility weights of severe lower respiratory tract infection ^28,29^ and the estimates of pricing models for COVID-19 treatments published by the Institute for Clinical and Economic Review ^23^ (Appendix 1.6).

We assumed a discount rate of 3% (0-6%) annually for both cost and quality-adjusted life-years (QALYs). We calculated the incremental costs and incremental QALYs for booster vaccination strategy compared with no booster (‘baseline’). The incremental cost-effectiveness ratio (ICER) was defined as the incremental cost per QALY gained. We used a cost-effectiveness threshold of ICER<$50,000. We conducted additional economic evaluations by calculating the benefit-cost ratio, cost/death saved, and net monetary benefit.

### Sensitivity analysis

We performed a univariate sensitivity analysis to examine the impact of model parameters within their respective ranges on the ICER to identify the most sensitive parameters and visualised the results using tornado diagrams. In addition, we conducted a probabilistic sensitivity analysis (PAS) based on 100,000 simulations to determine the probability of the booster strategy being cost-effective (**Figure 1**). The distributions of all model parameters were provided in *Appendix 1*.*7*. We conducted a 2-way sensitivity analysis to examine the impact of various combinations of vaccine efficacies, vaccination and direct medical cost on booster cost-effectiveness (**Figure 2**). By varying the population incidences between 0-50/100,000 person-day, we conducted the additional PSA (1,000 simulations for each population incidence) and presented the probabilities of the booster strategy being cost-effective and cost-saving (**Figure 3**). We further investigated the probabilities of being cost-effective and cost-saving under scenarios of less efficacious vaccines and boosters, where their efficacies for protection against infection and severe COVID-19 were reduced by 10%, 30%, and 50%, respectively. We estimated the threshold of population incidence for cost-effectiveness when the probabilities passed 50% for each scenario.

**Figure 1.**
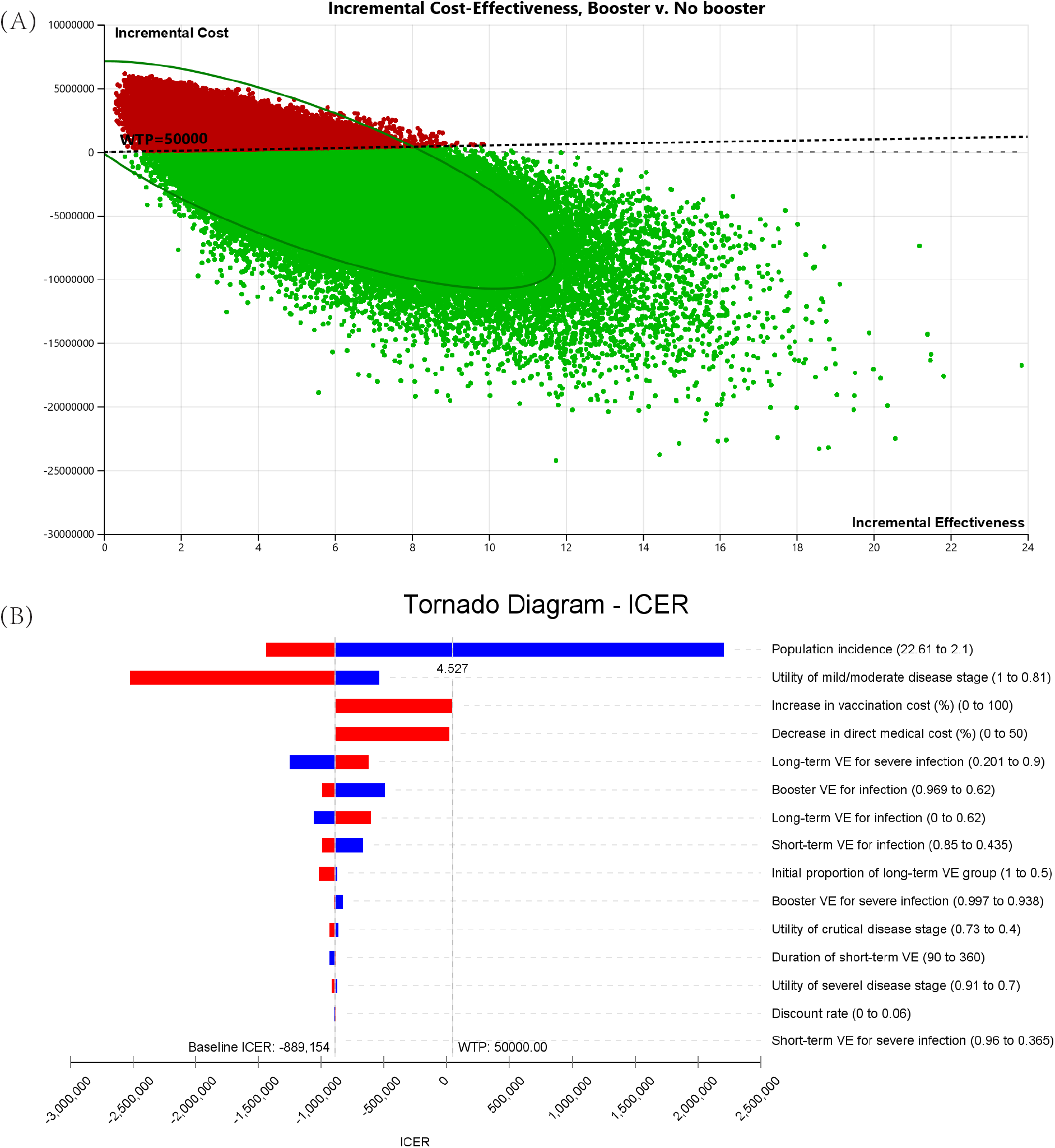
The cost-effectiveness analysis of Pfizer booster vaccination strategy. (A) the result of probabilistic sensitivity analysis based on 100,000 simulations (67.37% of being cost-effective, including 64.95% of being cost-saving); (B) tornado plot of one-way sensitivity analyses. A horizontal bar was generated for each parameter analyses. The width of the bar indicates the potential effect of the associated parameter on the ICER when the parameter is changed within its range (as shown in Table S1). The red part of each bar indicates high values of input parameter ranges, while the blue part indicates low values. The dotted vertical line represents the threshold of willingness-to-pay (WTP) of the baseline.

**Figure 2.**
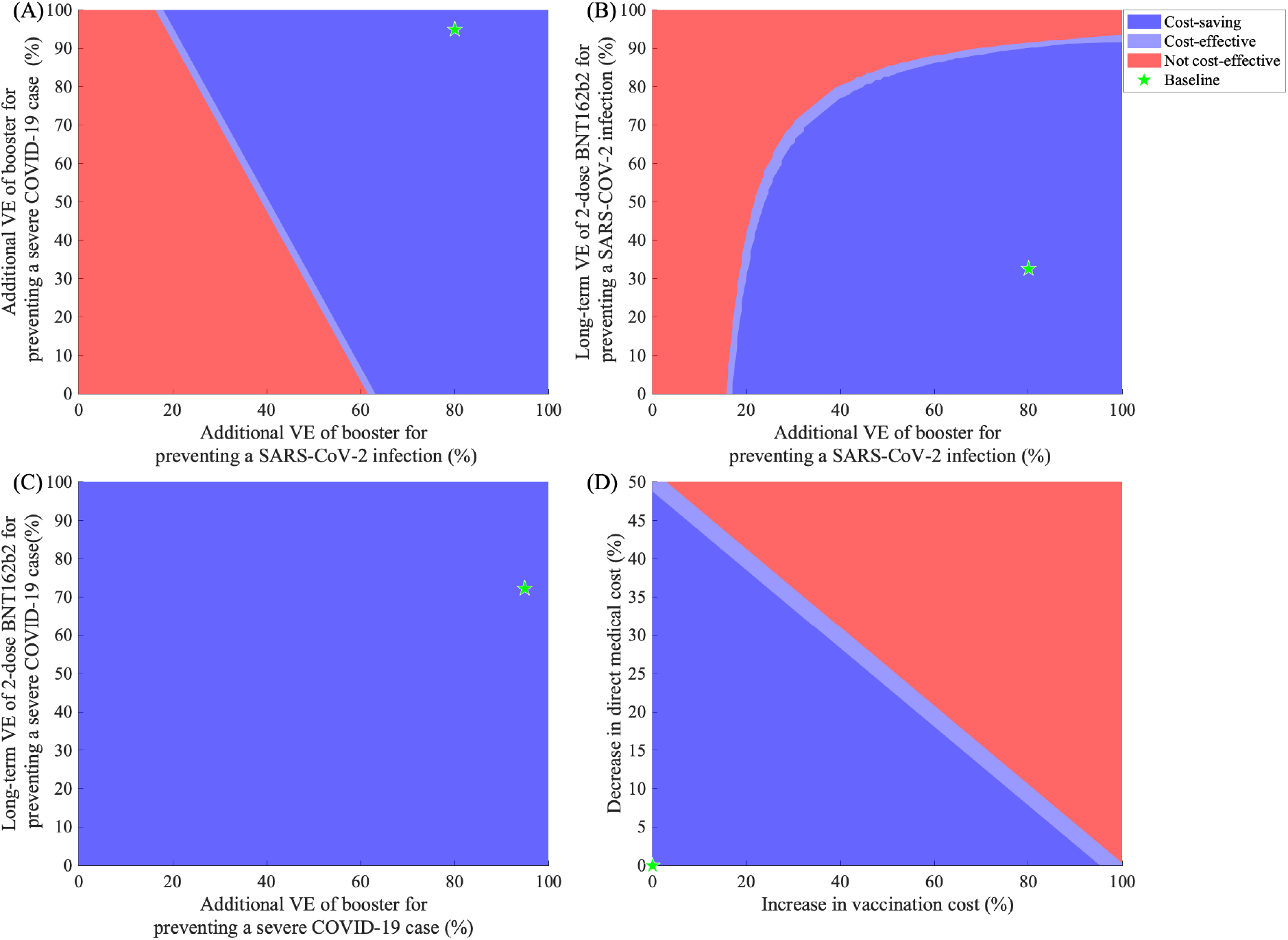
The result of 2-Way sensitivity analysis of Pfizer booster vaccination strategy. (A) Additional VE of booster for preventing a SARS-COV-2 infection and for preventing a severe COVID-19 case; (B) Additional VE of booster and Long-term VE of 2-dose BNT162b2 for preventing a SARS-COV-2 infection; (C) Additional VE of booster and Long-term VE of 2-dose BNT162b2 for preventing a severe COVID-19 case; (D) Vaccination and direct medical cost.

**Figure 3.**
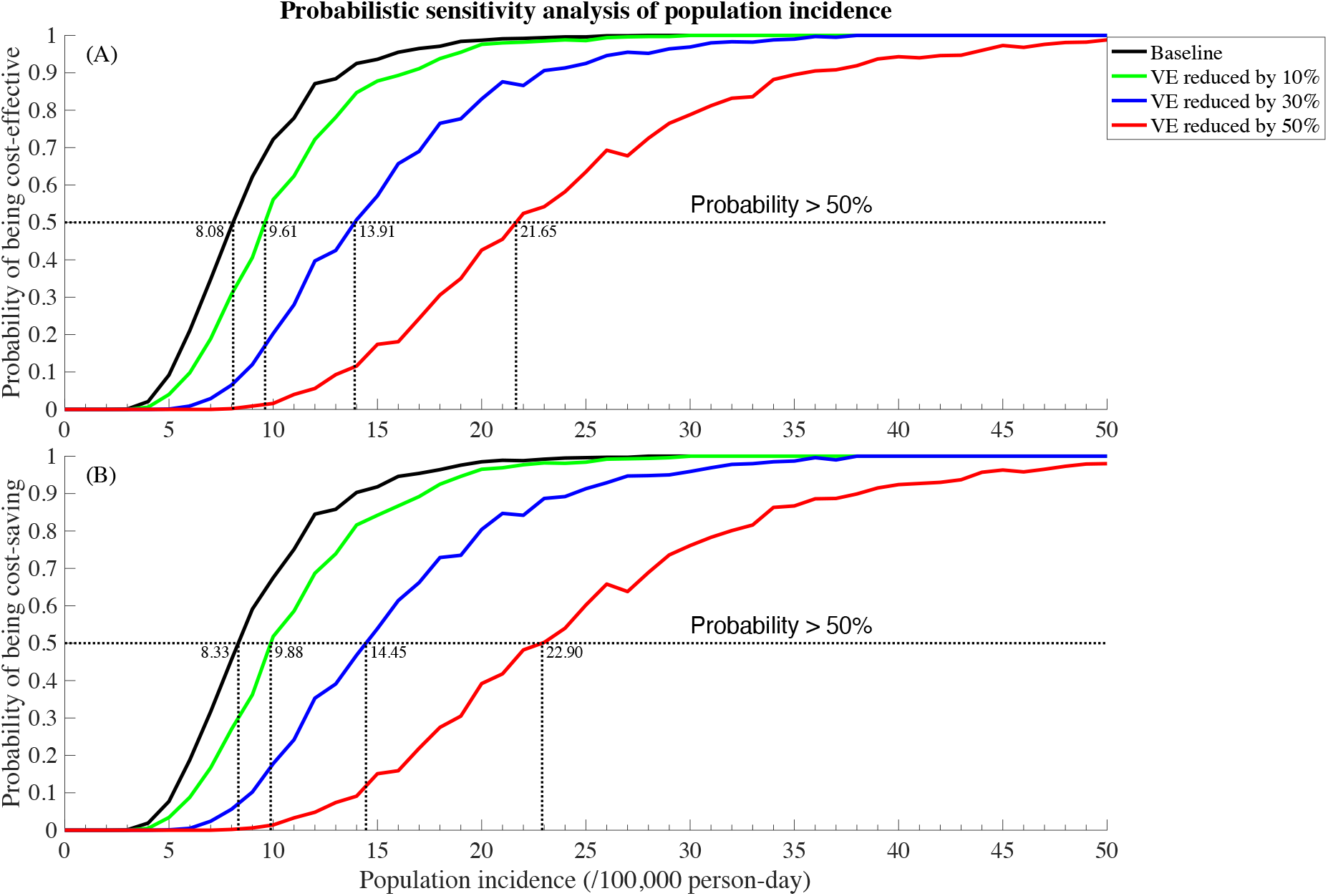
Probability and population incidence threshold of the Pfizer booster strategy being (a) cost-effective; (b) cost-saving with various vaccine (and corresponding booster) efficacies.

Existing evidence demonstrated that another mRNA COVID-19 vaccine Moderna mRNA-1273 had comparable or even higher efficacy than BNT162b2 ^30-32^, and the vaccine cost of Moderna mRNA-1273 was lower than that of BNT162b2 ^22^. Intuitively, Moderna mRNA-1273 would be more cost-effective than BNT162b2. In this study, we also conducted similar analyses for Moderna mRNA-1273 as a part of the sensitivity analysis (Appendix 1.8).

## Results

### Current BNT162b2 booster strategy is cost-saving in the US

We identified decremental costs and incremental QALYs for the BNT162b2 booster vaccination, compared with full-vaccination without boosters in a designated cohort of 100,000 older adults aged ≥65 years for 180 days. Overall, the booster strategy would incur an additional cost of $3,427,607 but save $6,691,935 dollars due to reduced direct medical care, corresponding to a benefit-cost ratio of 1.95. This suggested the booster strategy is a cost-saving one. Further, the strategy would result in a gain of 3.7 QALYs during the 180 days, and together with the monetary gain, it would amount to a net monetary benefit of $3,449,328. The strategy would prevent 3.8 COVID deaths, indicating a requirement of $904,382 to prevent one COVID death.

The probabilistic sensitivity analysis (PSA) based on 100,000 simulations demonstrated the probability of being cost-effective (including being cost-saving) with the current booster strategy was 67.37%, indicating a high chance of cost-effectiveness (**Figure 1A**). In contrast, the tornado diagram of univariate sensitivity analysis showed that varying any individual model parameter expect population incidence of COVID-19 at one time would not change the conclusion of cost-effectiveness of the booster strategy (**Figure 1B**). The population incidence of COVID-19 was the only factor that may alone alter the conclusion of cost-effectiveness of the booster strategy. We also noted that both the increase of vaccination cost and decrease in direct medical cost for COVID-19 treatment would reduce the cost-effectiveness of the booster strategy, but not sufficient to alter the conclusion individually.

### Impact of vaccine efficacies on booster cost-effectiveness

The 2-way sensitivity analysis showed that the booster strategy remained cost-effective at various combinations of vaccine efficacies. Figure 2A showed that if the booster provided 62% additional protection against the infection to fully-vaccinated older adults, it would be cost-effective even if the booster did not provide additional protection against the development of severe COVID-19 disease. Similarly, a combination of 50% additional protection against a SARS-COV-2 infection and 39% additional protection against severe COVID-19 would render the strategy cost-effective.

When comparing the protective efficacies of 2-dose vaccination against infection, with and without a booster, only when the long-term efficacy of a 2-dose vaccine program remained above 93% would a booster not confer sufficient additional protection to be cost-effective (Figure 2B). If the long-term efficacy against infection of a typical 2-dose vaccine only lay in the range of 30-40%, then a booster would only require to provide 18-20% additional protection to enable it to be cost-effective in those aged ≥65. On the contrary, when comparing the protective efficacies of a 2-dose vaccine against severe COVID-19 disease, the booster strategy would always be cost-effective (Figure 2C).

### Impact of vaccine and medical cost on booster cost-effectiveness

Doubling the vaccination cost or halving the direct medical cost for COVID-19 treatment alone would not change the cost-effectiveness status of the booster strategy (Figure 2D). However, certain combinations in the simultaneous changes of vaccination and medical cost, such as a 50% increase in vaccination cost and 26% reduction in direct medical cost for COVID-19 treatment, would render the booster strategy not cost-effective.

### Impact of population incidence and vaccine efficacies on booster cost-effectiveness

Figure 3 investigated the impact of varying population incidence (from 0 to 50/100,000 person-day) and declining vaccine and booster efficacies on the cost-effectiveness of the booster strategy. Our finding showed that for the current booster shots to be cost-effective (>50% chance), the population incidence of COVID-19 in the US aged ≥65 years needed to be ≥8.1/100,000 person-day. This threshold would increase with a decreasing vaccine and booster efficacy. For example, if the proactive efficacies against infection and severe COVID-19 disease by both the 2-dose vaccine and booster would reduce by 10% (as of BNT162b2), the population incidence threshold needed to be 9.6/100,000 person-day for the booster to be cost-effective. Further, if the vaccine and booster efficacies were reduced by 30-50% (a weak vaccine), the corresponding population incidence threshold would increase to 13.9 and 21.65/100,000 person-day. We also demonstrated similar results for the probability of being cost-saving (Figure 3B).

We also demonstrated similar key findings and conclusions regarding the cost-effectiveness of the Moderna mRNA-1273 booster as a part of sensitivity analysis (details in Appendix).

## Discussion

The study extensively evaluated the cost-effectiveness of a BNT162b2 booster strategy among older adults aged ≥65 years in the US. With an average population incidence of COVID-19 in older adults of 9.1/100,000 person-day, the probability that a booster strategy would be cost-effective is high (67%). In fact, our findings demonstrated that with every dollar of investment in the booster vaccination, the US government might save nearly two dollars due to fewer COVID-19 hospitalisation. Implementing the booster strategy in a cohort of 100,000 older adults would result in a net momentary benefit of $3.8 million in 180 days. Notably, the cost-effectiveness of the booster strategy is highly sensitive to the population incidence of COVID-19, with a threshold of 8.1/100,000 person-day being required to ensure the booster strategy to be cost-effective. This threshold will increase with a decrease in vaccine and booster efficacies.

Our study indicated that a COVID-19 booster strategy is likely to be cost-effective for older adults in the US. To the best of our knowledge, this is the first cost-effectiveness study of a COVID-19 booster strategy worldwide. We estimated that the booster strategy is cost-saving because the benefit of preventing one patient from being hospitalised and the subsequent needs of ICU and ventilation would outweigh the cost of delivering boosters to a large population of older adults. However, a combination of an increase in vaccine price (∼50%) and a decrease in direct medical cost (∼30%) may make boosters less cost-effective. As the demand for COVID-19 vaccines continue to surge worldwide and the increasing pressure faced by the US government to provide more vaccines to middle- and low-income countries, the vaccine price in the US may increase at some point. Further, the latest studies have documented significant development of antiviral drugs for COVID-19 treatment. Molnupiravir, the first oral medicine for the antiviral treatment of COVID-19, is highly effective in reducing viral loads in infected patients ^33^. Similarly, Paxlovid, another antiviral drug, has been shown to be 89% effective in patients at risk of serious illness ^34^. Since a novel and effective antiviral drug may potentially reduce the medical cost for treating patients with COVID-19, the booster strategy may become no longer cost-effective or even necessary in the future.

Our study indicates that the potential cost-effectiveness of the booster will reduce when the population incidence rate falls. In fact, the booster strategy will no longer be cost-effective if the population incidence in older adults reduces below 8.1/100,000 person-day. In a setting with an already high 2-dose vaccination coverage, the booster may further reduce the population incidence to below the threshold value in the elderly population and render it not cost-effective. However, a complete termination of the booster strategy may see a waning population immunity and subsequent rebound of the population incidence. If immunity falls after each booster, then a regular yearly vaccination program with further rounds of booster may be necessary to contain the epidemic to a low level.

Results from our sensitivity analyses suggest even in countries where vaccines with various efficacies are used a booster strategy may be cost-effective for older adults. For example, for vaccines whose 2-dose and booster efficacies were 10-50% lower than that of BNT162b2, a booster strategy would still be cost-effective as long as the COVID-19 incidence is greater than 21.7/100,000 person-day. Nevertheless, for countries with a low percentage of fully vaccinated population, it is essential to first achieve a high level of vaccination coverage before the booster strategy can be rolled-out.

Despite its cost-effectiveness, the booster strategy needs to be considered in the light of vaccine equity. So far, 14% of those ≥65 years of age in the US remain unvaccinated, and, therefore, efforts should be taken to help those unvaccinated or partially vaccinated receive their full vaccines in addition to providing boosters to those fully vaccinated. In addition, there exist stark racial and ethnic disparities in vaccination rate and COVID-19 disease outcomes in the US ^5^. African Americans reportedly have a lower vaccination rate and a greater disease burden of COVID-19 compared to Whites ^35^. Targeted interventions, such as culture-sensitive education and mass media campaigns that can improve the acceptance of COVID-19 vaccines among racial and ethnic minorities ^36,37^, should be implemented to reduce racial and ethnic disparities.

Our study has several limitations. First, our study used a decision-analytic Markov model and did not account for the dynamic changes of population incidence of COVID-19. Second, we estimated the efficacy of the BNT162b2 booster against the Delta variant based on a synthesis of evidence from real-world data rather than randomised controlled trials. We conducted various sensitivity analyses to account for uncertainty in real-world data and parameter biases. Third, we did not consider the other COVID-19 vaccines (e.g., Moderna or J&J/Janssen) in the US. While the other vaccines, particularly the Moderna vaccine, represents a substantial proportion of COVID-19 vaccines administered in the US, they differ from the BNT162b2 vaccines in efficacy and price. It may not be reasonable to combine different vaccines together. Nevertheless, the BNT162b2 vaccine comprises the largest proportion of all COVID-19 vaccines in the US; thus, the cost-effectiveness result of the BNT162b2 vaccines and boosters would be most policy-relevant. As the latest data showed that the Moderna vaccine could be more effective than the BNT162b2 vaccine in preventing hospitalisations ^31^, the cost-effectiveness of a booster strategy, in reality, is likely to be more favourable than what we estimated. Fourth, we assumed that the efficacy of COVID-19 vaccines begins to wane in 6 months after full vaccination. In reality, the efficacy of vaccines is more likely to gradually decline without a clear cut-off. This assumption may have led to an overestimate of vaccine efficacy in the short term and an underestimate in the long term. Fifth, we only evaluated the booster strategy for older adults. It is unclear whether the conclusion would be applicable to younger adults and children who have a lower vaccination rate but, at the same time, a lower risk of hospitalisation if diagnosed with COVID-19. This research question warrants further investigation. Finally, given limited public health resources and escalating health inequity during the pandemic, there is a need for more targeted, local-based vaccine and booster distribution strategies that can achieve a tradeoff between cost-effectiveness and equity. The design of such strategies, while beyond the scope of this work, will be critical in alleviating the burden of COVID-19, reducing health care costs, and achieving equity.

In conclusion, offering Pfizer-BioNTech booster shots to older adults aged ≥65 years in the US is likely to be cost-effective, but its cost-effectiveness is highly sensitive to the population incidence of COVID-19. Less efficacious vaccines and boosters may still be cost-effective in settings of high SARS-COV-2 transmission.

**Table 1.** The results of cost-effectiveness analysis of BNT162b2 booster vaccination for COVID-19 in elderly ≥65 years in US. (Reference scenario: elderly fully vaccinated with BNT162b2 but not booster; a cohort of 100,000 individuals over an evaluation period of 180 days)

|  | Full vaccination with BNT162b2 | Full vaccination with BNT162b2 + booster | Incremental benefits* |
| --- | --- | --- | --- |
| QALY | 48,908.4 | 48,912.1 | 3.7 |
| Uninfected individuals | 48,526.1 | 48,796.9 | -- |
| Infected individuals | 383.3 | 115.2 | -- |
| Costs, \$ | 8,901,608 | 5,637,280 | -\$3,264,328 |
| Vaccination cost | 0 | 3,427,607 | \$3,427,607 |
| Direct medical cost | 8,901,608 | 2,209,673 | -\$6,691,935 |
| Death cases | 4.66 | 0.87 | 3.79 |
| ICER | -- | -- | Cost saving |
| Benefit-cost ratio | -- | -- | 1.95 |
| Cost/death prevented, \$ | -- | -- | \$904,382 |
| Net monetary benefit, \$ | -- | -- | \$3,449,328 |

\* Incremental benefits = difference between the booster and the reference scenarios.
Benefit-cost ratio: each dollar invested in vaccination will save 1.95 dollars of direct medical cost.
Cost/death prevented: every 904,382-dollar invested in vaccination will prevent one death.
Net monetary benefit (NMB) is calculated as (incremental benefit x threshold) – incremental cost.

## Data Availability

Not applicable

## Declarations

### Ethics approval and consent to participate

Not applicable

### Authors’ contributions

LZ conceived the study. LZ, YL, MS and RL designed and constructed the model. RL performed the modelled analyses, graphed and interpreted the results. RL, HL, ZZ, LX and XL contributed to the collection of data and model parameters. RL, LZ, YL and HL drafted the manuscript. LZ, YL and MS critically revised the manuscript. All authors reviewed the manuscript and approved the final version.

