## Appendix for "Cost-effectiveness analysis of BNT162b2 COVID-19 booster vaccination in the United States"

This supplementary document describes in detail the model construction and estimation of parameters presented in the main text.

**1.1 Health stages**

We developed a decision-analytic Markov model to simulate the disease progression of SARS-CoV-2 infection in a designated initial cohort of 100,000 individuals aged ≥65 years over a period of 180 days. The model consisted of 9 health states depicting varied disease progression of COVID-19 **(Figure S1).** Existing evidence indicated that the vaccine efficacy (VE) of Pfizer-BioNTech BNT162b2 would gradually wane after six months ^1-3^. We defined the nine diseases states as follows.

After being infected by SARS-CoV-2 strains, a fully vaccinated individual may progress through the following health states.

- Short-term VE: the vaccine efficacy from 2 weeks to 6 months after the 2nd dose of vaccines
- Long-term VE: the vaccine efficacy six months beyond the 2nd dose
- Incubation: cases prior to symptom onset
- Asymptomatic: cases who never developed symptoms ever throughout the course of their disease
- Mild/moderate: cases without pneumonia and cases with mild pneumonia
- Severe: cases who developed dyspnoea and/or hypoxemia and managed in a hospital but not requiring intensive care unit (ICU)
- Critical: cases who developed respiratory failure, and/or septic shock, and/or multiple organ dysfunction/failure and managed in an ICU; some of them who recuperated from critical disease need to go through the recuperation stage-remaining in the hospital or other health care facility
- Recovered: cases who recovered from infection stages; we assumed that a recovered individual could not be reinfected for the next 180 days (similar to the short-term vaccine protection)
- Death: COVID-19 related death

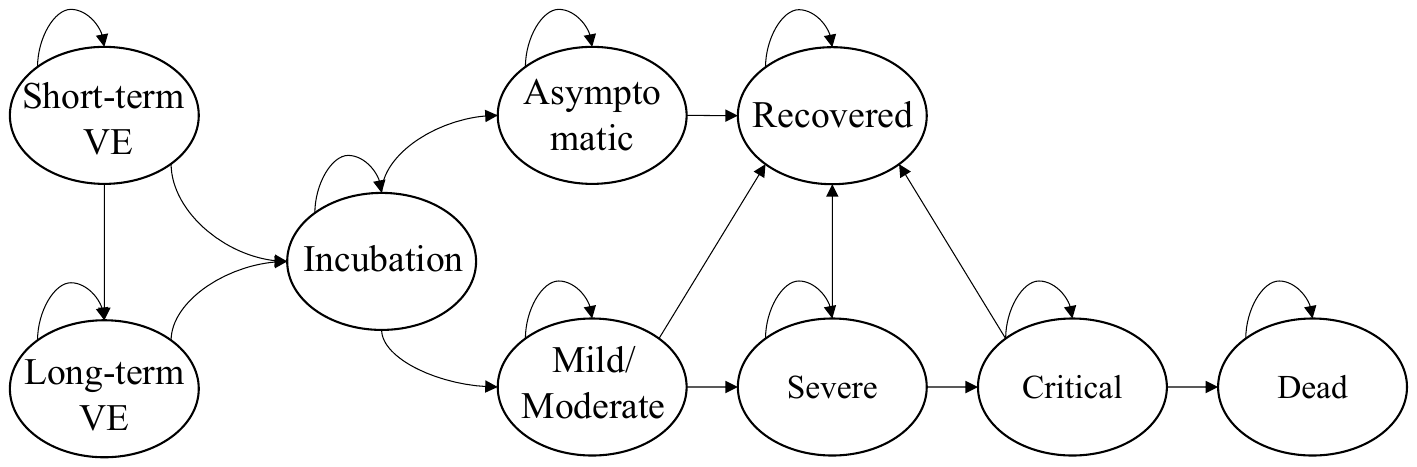

**Figure S1.** Markov model of COVID-19 natural history in the vaccinated population

**1.2 Vaccine efficacy**

We collected information on the vaccine efficacy of BNT162b2 for SARS-CoV-2 (Delta variant) infection in older adults aged ≥65 years based on an ongoing systematic review conducted by The International Vaccine Access Center ^4^. We extracted ten studies from the review and included one additional study supplemented by a PubMed search according to the search strategy (BNT162b2 [Title/Abstract]) AND (Delta [Title/ Abstract]) on 28th October 2021.

Of the 11 collected studies, seven were test-negative case-control studies (Odds ratio, OR, as risk measurement), and four were retrospective cohort studies (Hazard ratio, HR, as risk measurement). The test-negative case-control studies compared the risk of infection and severe COVID-19 disease between older adults who were fully vaccinated with those who received no vaccination (**Table S2**). We conducted a meta-analysis to obtain a pooled OR, grouped by vaccination period and outcome measure (**Figure S2**). We then calculated the vaccine efficacy using the formula: (1–OR) multiplied by 100% ^2^. Due to the small number of eligible retrospective cohort studies, we calculated a simple arithmetic average for the VE. We then further calculated the overall VE by averaging the efficacies from the two groups and adapting their lowest and highest bounds.

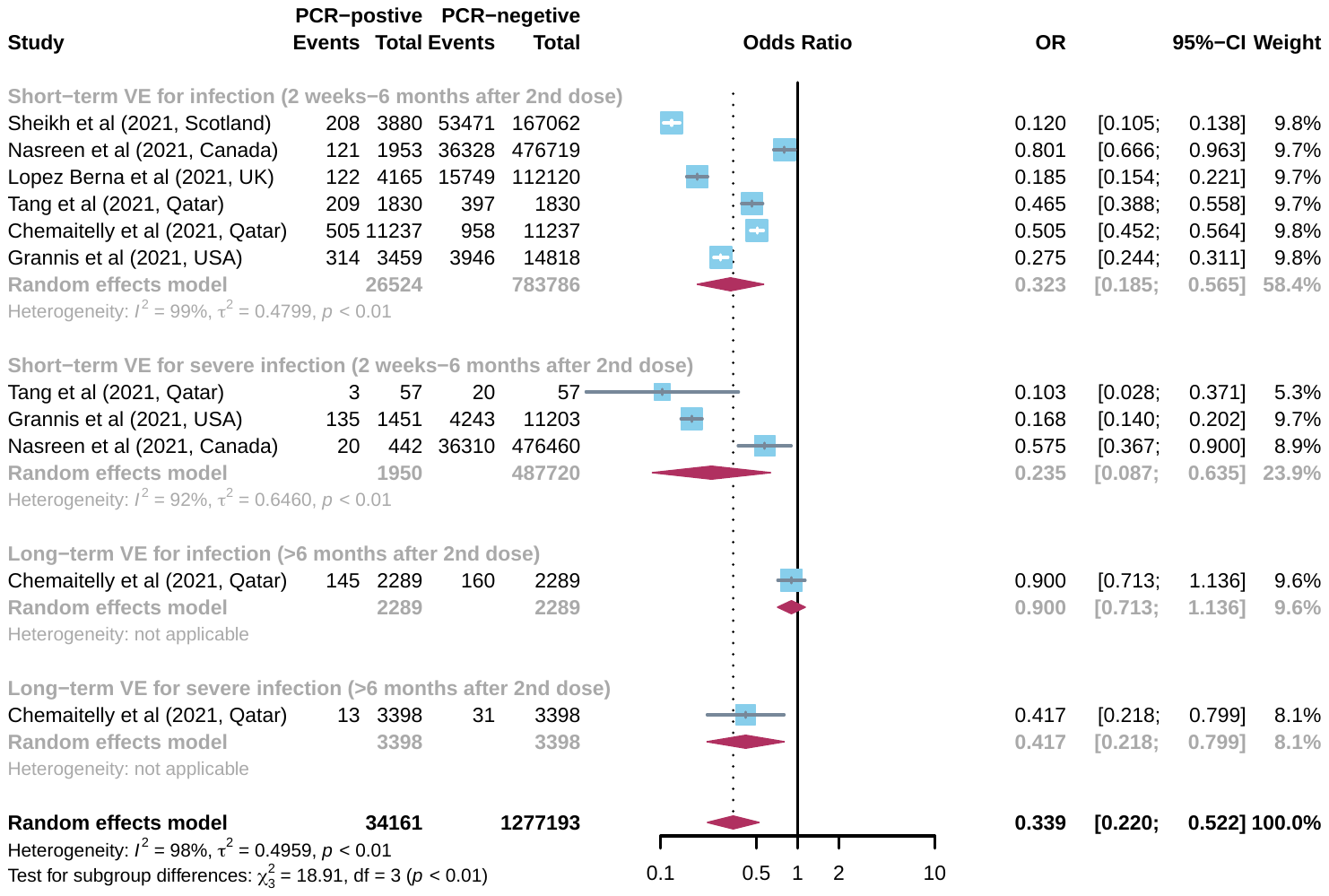

**Figure S2**. Forest plot for the pooled Odds Ratio of fully vaccinated BNT162b2 individuals against SARS-CoV-2 variant (Delta).

| Variables | Test-negative case-control designs | Retrospective cohort  designs | **Average VE** |
| --- | --- | --- | --- |
| Short-term VE for preventing Delta infection (%) | 67.0 (43.5, 81.5) | 76.0 (58.0, 85.0) | **71.5 (43.5, 85.0)** |
| Short-term VE for preventing severe COVID-19 disease (%) | 76.5 (36.5, 91.3) | 90.3 (77.0, 96.0) | **83.4 (36.5, 96.0)** |
| Long-term VE for preventing Delta infection (%) | 10.0 (-13.6, 28.7) | 54.9 (48.0, 62.0) | **32.5 (0.00, 62.0)** |
| Loge-term VE for preventing severe COVID-19 disease (%) | 58.3 (20.1, 78.2) | 86.0 (82.0, 90.0) | **72.2 (20.1, 90.0)** |

Based on the retrospective cohort study conducted in Israel for older aged ≥60 years ^5^ (we assume the same association applied to those aged ≥65 years), we estimated the additional vaccine efficacy of the booster strategy in fully-vaccinated individuals compared with those without a booster. The formula was: (1–1/HR) ⅹ100% ^6^.

| **Variables** | **Hazard ratio (HR)** | **VE (%)** |
| --- | --- | --- |
| Additional booster VE for preventing infection, compared with full vaccination without a booster | 11.4 (10.4, 12.3) | 80.1 (62.0, 91.9) |
| Additional booster VE for preventing severe infection, compared with full vaccination without a booster | 19.5 (12.9, 29.5) | 94.9 (92.2, 96.6) |
| Booster VE for preventing infection, compared with no vaccination | **—** | **86.6 (62.0, 96.9)** |
| Booster VE for preventing severe infection, compared with no vaccination | **—** | **98.6 (93.8, 99.7)** |

**1.3 Distribution of clinical disease stages of various vaccination status**

We collected the distribution of clinical disease stages after being infected by SARS-CoV-2 strains in unvaccinated group from published literature ^7-10^. Based on the VE of short-term, long-term and booster, we developed a mathematical model to estimate the similar distributions in vaccinated group (**Figure S3**). In the model, we calculated the numerator and denominator of the proportion of severe infections in vaccinated patients ($P_{vs}$) by three factors, which are the proportion of severe infections in unvaccinated infections ($P_{s}$), the vaccine efficacy for preventing a SARS-COV-2 infection ($V_{i}$) and for preventing a severe COVID-19 case ($V_{s}$).

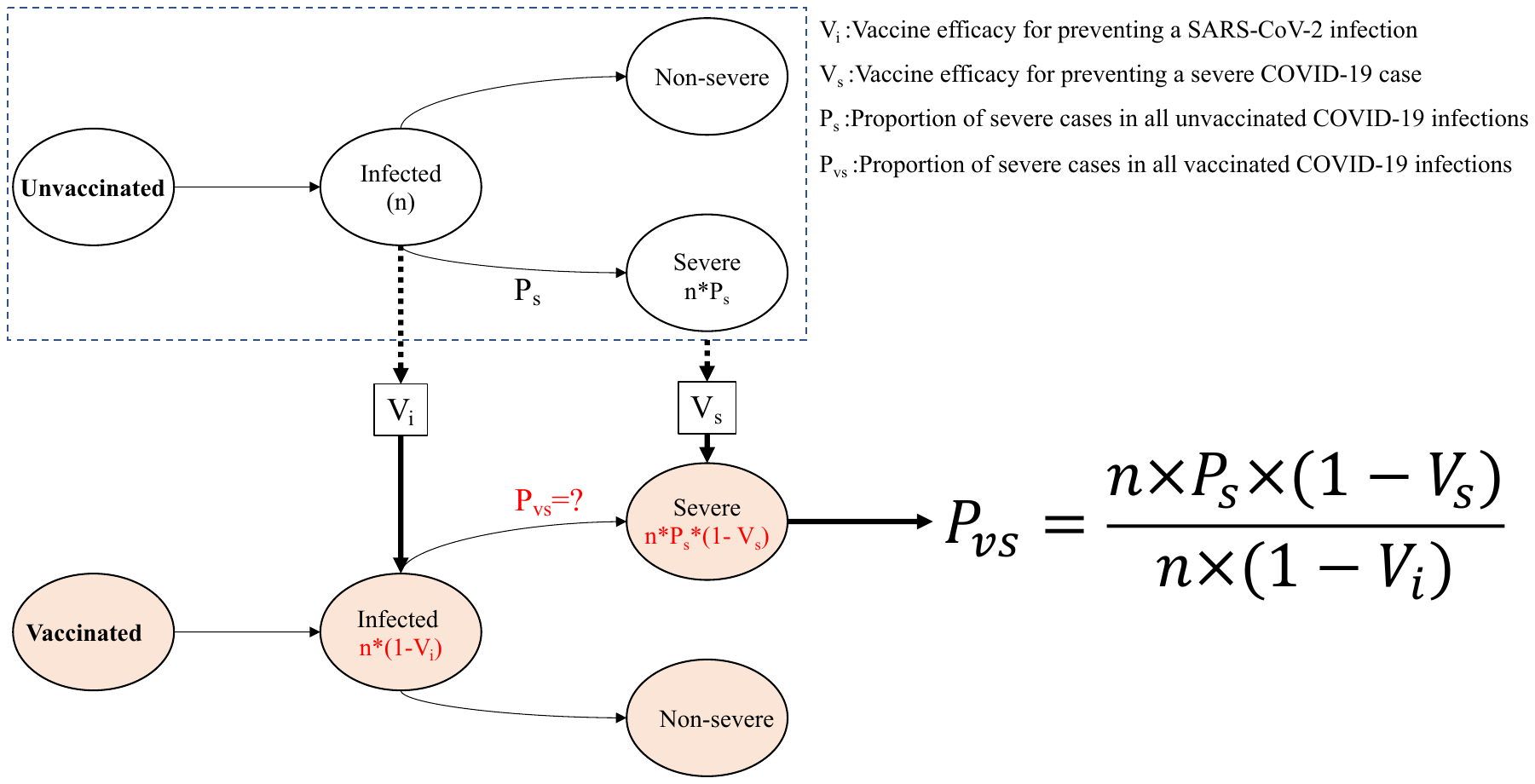

**Figure S3**. The model flowchart to estimate the proportion of severe cases in all COVID-19 infections who have got fully vaccinated.

Then, we could obtain the proportion of the severe cases (including severe and critical) after COVID-19 infection in various vaccination status. We assumed the proportion of critical cases in severe infections was fixed both in unvaccinated and vaccinated groups. Thus, we could calculate the proportion of severe and critical clinical outcome after COVID-19 infections in varied vaccinated group, according to that in unvaccinated group. Similarly, we calculated the proportion of asymptomatic and mild/moderate clinical outcome after COVID-19 infections in varied vaccinated group. The details of the distribution are list as follow.

| **Clinical outcomes** | **The distribution of unvaccinated group** | **The distribution of short-term VE group** | **The distribution of long-term VE group** | **The distribution of booster VE group** |
| --- | --- | --- | --- | --- |
| Asymptomatic | 17.10% | 17.60% | 17.80% | 18.17% |
| Mild/Moderate | 76.37% | 78.58% | 79.50% | 81.15% |
| Severe | 1.400% | 0.815% | 0.577% | 0.146% |
| Critical | 5.160% | 3.005% | 2.125% | 0.539% |
| Critical (recover) | 4.363% | 2.541% | 1.797% | 0.456% |
| Critical (die) | 0.797% | 0.464% | 0.328% | 0.083% |

**1.4 Population incidence of COVID-19 Delta strain.**

The Centers for Disease Control and Prevention (CDC) reported the weekly COVID-19 new infections and death cases per 100,000 population by age in the US ^11^. We estimated the population incidence of COVID-19 in US older adults to be 9.12/100,000 person-day by averaging out the published data (of US CDC) over the last 180 days before the approval of a booster shot on 22^nd^ September 2021. We adopt the highest and the lowest incidence (2.10-22.61/00,000 person-day) during the same period as its range for sensitivity analysis.

During the last 180 days before 22^nd^ September 2021, the population incidence of COVID-19 in the US consists of three vaccination status of unvaccinated, short-term VE and long-term VE. The contribution of each status for population incidence are dependent on the population proportion and its infection risk (1-VE) of the status. Based on the average population proportion of three status (35.55%, 13.26%, 51.19%)^12^ and its various VE, we calculated the population incidence in unvaccinated, short-term VE and long-term VE group are 15.43/100,000, 4.40/100,000, and 10.42/100,000 person-day, respectively. Then, we calculated booster VE population incidence are 2.07/100,000 person-day.

**1.5 Direct medical cost**

We collected the total direct medical costs from hospitalisation at various COVID-19 clinical stages from the Projected Economic Impact Report of the US Healthcare System and Health System Tracker^13,14^. We assumed all the severe and critical COVID-19 infections would be admitted to hospitals and estimated that 11.05% of the Mild/Moderate COVID-19 infection would be admitted to the hospital. We estimated the overall cost of hospitalisation at various clinical disease stages. We then calculated the corresponding daily cost by dividing the total hospitalisation cost by the duration of the clinical stages (**Table S1**).

| Clinical outcomes | Distribution of clinical disease stages | Proportion of hospitalisation at various clinical disease stages | Total cost of hospitalisation at various clinical disease stages, $ | Total cost at various clinical disease stages, $ | Duration of the clinical disease stages (days) | Medical cost/day $ |
| --- | --- | --- | --- | --- | --- | --- |
| Asymptomatic | 17.10% | 0% | 0 | 0 | 0 | 0 |
| Mild/Moderate | 76.37% | 11.05%* | 42,486 | 4695 | 10 | 470 |
| Severe | 1.400% | 100% | 53,268 | 53,268 | 6.5+10.5 | 4782 |
| Critical (die) | 4.363% | 100% | 74,310 | 74,310 | 3+7.1+11.9 | 3273 |
| Critical (recover) | 0.797% | 100% | — | 76,986 | 3+7.1+11.9+5.7 | **—** |
| * The proportion of hospitalisation of Mild/Moderate infection was calculated by the total hospitalisation rate of COVID-19 infection (~15%) ^13^. | | | | | | |

**1.6 Health state utilities**

We collected utility scores for COVID-19 patients from the disutility weights of severe lower respiratory tract infection ^15,16^ and the estimates of pricing models for COVID-19 treatments published by the Institute for Clinical and Economic Review ^17^. We calculated the average utility from two sources and adopted their lowest and highest bounds.

| Health states | Disability weight 1 ^15,16^ | Disability weight 2 ^17^ | **Average utility** |
| --- | --- | --- | --- |
| In asymptomatic state | — | — | **1** |
| In mild/moderate state | — | 0.19 | **0.905 (0.810, 1.000)** |
| In severe state | 0.13 (0.09, 0.19) | 0.30 | **0.785 (0.700, 0.910)** |
| In critical state | 0.41 (0.27, 0.56) | 0.50-0.60 | **0.520 (0.400, 0.730)** |
| In recuperable state | — | — | **0.905 (0.810, 1.000)** |
| In dead state | 0 | — | **0** |

**1.7 Sensitivity analyses**

We extensively explored the impact of model parameters on the baseline results with univariate, 2-way and probabilistic sensitivity analyses (PSA). Univariate sensitivity analysis is for each of 15 parameters and varying one parameter at one time in their range. 2-way sensitivity analyses focuses on the combination of four couple parameters in more extensive range (additional booster VE against infection and severe disease, additional booster and long-term VE against infection and severe disease, additional booster and long-term VE against severe disease, vaccination costs and direct medical cost). PSA allows variation of all 15 parameters together at one time in their distribution.

| **No** | **Name** | **Distribution** | **Reference** |
| --- | --- | --- | --- |
| 1 | Duration of short-term VE | Uniform (90, 360) | Assumed and ^1-3^ |
| 2 | Initial proportion of Long-term VE group | Uniform (0.5, 1.0) | Assumed and ^18^ |
| 3 | Population incidence | Triangular (2.10, 9.12, 22.61) | Appendix 1.4 |
| 4 | Short-term VE for preventing Delta infection | Triangular (0.435, 0.715, 0.850) | Appendix 1.2 |
| 5 | Short-term VE for preventing severe COVID-19 disease | Triangular (0.365, 0.834, 0.960) | Appendix 1.2 |
| 6 | Long-term VE for preventing Delta infection | Triangular (0, 0.325, 0.620) | Appendix 1.2 |
| 7 | Loge-term VE for preventing severe COVID-19 disease | Triangular (0.201, 0.722, 0.900) | Appendix 1.2 |
| 8 | Booster VE for preventing infection, compared with no vaccinated | Triangular (0.620, 0.866, 0.969) | Appendix 1.2 |
| 9 | Booster VE for preventing severe infection, compared with no vaccinated | Triangular (0.938, 0.986, 0.997) | Appendix 1.2 |
| 10 | Decrease in direct medical cost (%) | Uniform (0, 50) | Assumed |
| 11 | Increase in vaccination cost (%) | Uniform (0, 100) | Assumed |
| 12 | Utility of mild/ moderate disease stage | Triangular (0.810, 0.905, 0.100) | Appendix 1.6 |
| 13 | Utility of severe disease stage | Triangular (0.700, 0.785, 0.910) | Appendix 1.6 |
| 14 | Utility of critical disease stage | Triangular (0.400, 0.520, 0.730) | Appendix 1.6 |
| 15 | Discount rate | Uniform (0.00, 0.06) | ^19^ |

**1.8 The cost-effectiveness of Moderna booster vaccination strategy**

We further evaluated the cost-effectiveness of Moderna booster strategy. Existing evidence demonstrated that another mRNA COVID-19 vaccine Moderna mRNA-1273 had comparable or even higher efficacy than BNT162b2^20-22^. Thus, we assumed that all the VE of Moderna (short-term, long-term, booster) are higher by 5% than those of Pfizer-BioNTech. In addition, a single vaccine dose of Moderna is $15.0^23^, lower than Pfizer-BioNTech ($19.5).

Similarly, we also identified the decremental costs and incremental QALYs for the Moderna booster vaccination strategy, compared with full-vaccination without boosters in a designated cohort of 100,000 older adults aged ≥65 years for 180 days. Overall, the booster strategy would incur an additional cost of $3,006,491, but save $6,837,746 dollars due to reduced direct medical care, corresponding to a benefit-cost ratio of 2.27. This suggested the booster strategy is a cost-saving one. Further, the strategy would result in a gain of 3.7 QALYs during the 180 days, and together with the monetary gain would amount to a net monetary benefit of $3,191,491. The strategy would prevent 3.6 COVID deaths, indicating a requirement of $836,277 to prevent one COVID death.

**Table S4.** The results of cost-effectiveness analysis of Moderna booster vaccination for COVID-19 in elderly ≥65 years in US.

|  | Full vaccination with Moderna | Full vaccination with Moderna + booster | Incremental benefits* |
| --- | --- | --- | --- |
| QALY | 48,908.6 | 48,912.3 | 3.7 |
| Uninfected individuals | 48,527.0 | 48,814.5 | -- |
| Infected individuals | 381.6 | 97.8 | -- |
| Costs, $ | 8,575,190 | 4,743,935 | -$3,831,255 |
| Vaccination cost | 0 | 3,006,491 | 3,006,491 |
| Direct medical cost | 8,575,190 | 1,737,444 | -$6,837,746 |
| Death cases | 4.12 | 0.52 |  |
| ICER | -- | -- | Cost saving |
| Benefit-cost ratio | -- | -- | 2.27 |
| Cost/death prevented, $ | -- | -- | 836,277 |
| Net monetary benefit, $ | -- | -- | 3,191,491 |
| * Incremental benefits = difference between the booster and the reference scenarios.  Benefit-cost ratio: each dollar invested in vaccination will save 2.27 dollars of direct medical cost.  Cost/death prevented: every 836,277-dollar invested in vaccination will prevent one death.  Net monetary benefit (NMB) is calculated as (incremental benefit x threshold) – incremental cost. | | | |

The probabilistic sensitivity analysis (PSA) demonstrated the probability of being cost-effective by varying all model parameters within its range simultaneously (**Figure S4A**). Among the 100,000 simulations, the probability of being cost-effective (including being cost-saving) with the current booster strategy was 75.97%, indicating a high chance of cost-effectiveness. In contrast, the tornado diagram of univariate sensitivity analysis showed that varying any single model parameters expect population incidence of COVID-19 at one time would not change the conclusion of cost-effectiveness of the booster strategy (**Figure S4B**). In fact, the population incidence of COVID-19 was the only factor that may alone alter the conclusion of cost-effectiveness of the booster strategy. We also noted that both of the increase of vaccination cost and decrease in direct medical cost for COVID-19 treatment would reduce the cost-effectiveness of the booster strategy, but not sufficient to alter the conclusion individually.

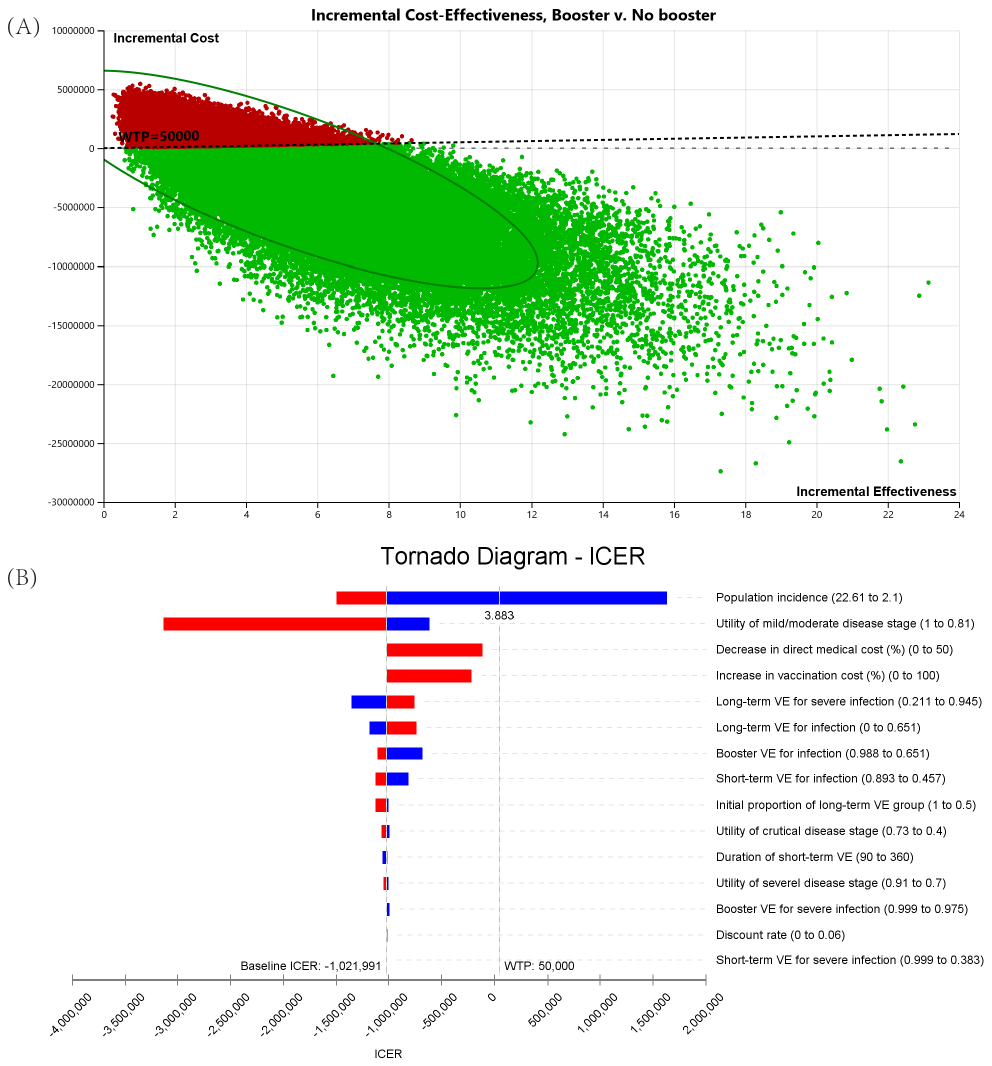

**Figure S4** The cost-effectiveness analysis of Moderna booster vaccination strategy (A) the result of probabilistic sensitivity analysis based on 100,000 simulations (75.97% of being cost-effective, including 73.82% of being cost-saving); (B) tornado plot of one-way sensitivity analyses. A horizontal bar was generated for each parameter analyses. The width of the bar indicates the potential effect of the associated parameter on the ICER when the parameter is changed within its range (as shown in Table S1). The red part of each bar indicates high values of input parameter ranges, while the blue part indicates low values. The dotted vertical line represents the threshold of willingness-to-pay (WTP) of the base-case.

The 2-way sensitivity aanalyses showed that the booster strategy remained cost-effective at various combinations of vaccine efficacies. **Figure S5A** showed that if the booster would offer 53% additional protection against the infection to fully-vaccinated individual's aged ≥65 years with a long-tern waning immunity, it would be already cost-effective even if the booster did not provide additional protection against the development of severe COVID-19. Similarly, a combination of 50% additional protection against a SARS-COV-2 infection and 33% additional protection against severe COVID-19 would render the strategy cost-effectiveness. When comparing the protective efficacies of 2-dose vaccination against an infection, with and without a booster, **Figure S5B** demonstrated that only when the long-term efficacy of a 2-dose vaccine program remained above 95%, did a booster would not confer sufficient additional protection to be cost-effective. If the long-term efficacy of a 2-dose vaccine only lied in the range of 30-40%, then a booster only would only be required to provide an 13-14% additional protection to enable it to be cost-effective in those ≥65. On the contrary, when comparing the protective efficacies of 2-dose vaccine against severe COVID-19, the booster strategy would always be cost-effective (**Figure S5C**). In all cases, the booster conferred very high efficacies against both infection and severe disease and would be cost-saving.

Similarly, doubling the vaccination cost or halving the direct medical cost for COVID-19 treatment alone would not change the cost-effectiveness status of the booster strategy (**Figure S5D**). However, certain combinations in the simultaneous changes of vaccination and medical cost, such as 50% increase in vaccination cost and 37% reduction in direct medical cost for COVID-19 treatment, would render the booster strategy not cost-effective.

**Figure S5** The result of 2-Way sensitivity analysis of Moderna booster vaccination strategy. (A) Additional VE of mRNA-1273 booster for preventing a SARS-COV-2 infection and for preventing a severe COVID-19 case; (B) Additional VE of booster and Long-term VE of 2-dose mRNA-1273 for preventing a SARS-COV-2 infection; (C) Additional VE of booster and Long-term VE of 2-dose mRNA-1273 for preventing a severe COVID-19 case; (D) Vaccination and direct medical cost.

**Table S1 Model parameters for cost-effectiveness analysis of COVID-19 BNT162b2 booster vaccination for ≥65ys population in the US**

| **Model parameters** | **Elderly received the 2^nd^ dose of BNT162b2 for 14-180 days** | **Elderly received the 2^nd^ dose of BNT162b2 for >180 days** | **Fully vaccinated elderly received BNT162b2 booster** | **Reference** |
| --- | --- | --- | --- | --- |
| **Epidemiological parameters** |  | | |  |
| Population incidence of COVID-19 before the approval of booster shots (cases/100,000 person-day) | 4.40 (1.01, 10.91) | 10.42 (2.40, 25.83) | 2.07 (0.48, 5.17) | Estimated based on ^4,5,24^, details in Appendix 1.4 |
| Duration of clinical disease stages of COVID-19 (days) | | | | ^25-27^ |
| Incubation | 5.2 (4.1, 7.0) | | |  |
| Asymptomatic infection | 6.0 (2.0, 12.0) | | |  |
| Mild/Moderate illness | 10.0 | | |  |
| Severe illness |  | | |  |
| In mild/moderate state | 6.5 | | |  |
| In severe state | 10.5 | | |  |
| Critical (recover) illness |  | | |  |
| In mild/moderate state | 3.0 | | |  |
| In severe state | 7.1 | | |  |
| In critical state | 11.9 | | |  |
| In recuperable state | 5.7 | | |  |
| Critical (die) illness |  | | |  |
| In mild/moderate state | 3.0 | | |  |
| In severe state | 7.1 | | |  |
| In critical state | 11.9 | | |  |
| **Costing parameters** |  | | |  |
| Cost of COVID-19 PCR test, $ | 51 | | | ^28^ |
| Cost of COVID-19 vaccine booster, $ | 0 | 0 | 19.50 | ^23^ |
| Cost of vaccine administration, $ | 0 | 0 | 17.10 | ^17,29^ |
| Direct medical cost of clinical disease outcomes of COVID-19 infection, $ | | | | ^13,14^ |
| Asymptomatic infection | 0 | | |  |
| Mild/Moderate illness | 4695 | | |  |
| Severe illness | 53,268 | | |  |
| Critical (recover) illness | 76,986 | | |  |
| Critical (die) illness | 74,310 | | |  |
| **Life quality parameters** |  | | |  |
| Utility weight of clinical disease progression severity | | | | ^15-17^ |
| In asymptomatic state | 1 | | |  |
| In mild/moderate state | 0.905 (0.810, 1.000) | | |  |
| In severe state | 0.785 (0.700, 0.910) | | |  |
| In critical state | 0.520 (0.400, 0.730) | | |  |
| In recuperable state | 0.905 (0.810, 1.000) | | |  |
| In dead state | 0 | | |  |
| Discount rate, per year | 3% (0%, 6%) | | | ^19^ |

**Tabel S2 Fully vaccinated BNT162b2 efficacy (VE) for SARS-CoV-2 variant (Delta) infection in elderly aged ≥ 65 years in studies with a test-negative case-control design**

| No | First author | Country | Duration | Cases (PCR-positive) | | Control (PCR-negative) | | VE(%) | Note |
| --- | --- | --- | --- | --- | --- | --- | --- | --- | --- |
|  |  |  |  | Vaccinated | Unvaccinated | Vaccinated | Unvaccinated |  |  |
| **Short-term VE for preventing Delta infection (2 weeks-6 months after 2nd dose)** | | | | | | | | | |
| 42 | Sheikh et al ^30^ | Scotland | Apr 1, 2021-Jun 1, 2021 | 208 | 3,672 | 53,471 | 113591 | 79.0 (75.0, 82.0) | 14-140 days, all age |
| 49 | Nasreen et al ^31^ | Canada | Apr 5, 2021-Aug 3, 2021 | 121 | 1,832 | 36,328 | 440,391 | — | >14 days, all age |
| 62 | Lopez Berna et al ^32^ | UK | Dec 1, 2020-May 30, 2021 | 122 | 4,043 | 15,749 | 96,371 | 88.0 (85.3, 90.1) | 14-109 days, all age |
| 74 | Tang et al ^33^ | Qatar | Dec 21, 2020-Jul 21, 2021 | 209 | 1,621 | 397 | 1,433 | 53.5 (43.9, 61.4) | 14-175 days, all age |
| 82 | Chemaitelly et al ^2^ | Qatar | Dec 21, 2020-Sep 5, 2021 | 505 | 10,732 | 958 | 10,279 | — | 1-6 months, all age |
| 92 | Grannis et al ^34^ | USA | Jun 1, 2020-Aug 31, 2021 | 314 | 3,145 | 3,946 | 10,872 | 77.0 (74.0, 80.0) | 14-180 days, all age |
| **Short-term VE for preventing severe COVID-19 disease (2 weeks-6 months after 2nd dose)** | | | | | | | | | |
| 74 | Tang et al ^33^ | Qatar | Dec 21, 2020-Jul 21, 2021 | 3 | 54 | 20 | 37 | 89.7 (61.0, 98.1) | 14-175 days, all age |
| 92 | Grannis et al ^34^ | USA | Jun 1, 2020-Aug 31, 2021 | 135 | 1,316 | 4,243 | 6,960 | 80.0 (73.0, 85.0) | 14-180 days, all age |
| 49 | Nasreen et al ^31^ | Canada | Apr 5, 2021-Aug 3, 2021 | 20 | 422 | 3,6310 | 440,150 | — | >14 days, all age |
| **Long-term VE for preventing Delta infection (>6 months after 2nd dose)** | | | | | | | | | |
| 82 | Chemaitelly et al ^2^ | Qatar | Dec 21, 2020-Sep 5, 2021 | 145 | 2,144 | 160 | 2,129 | 17.9 (-12.9, 40.3) | >6 months, all age |
| **Long-term VE for preventing severe COVID-19 disease (>6 months after 2nd dose)** | | | | | | | | | |
| 82 | Chemaitelly et al ^2^ | Qatar | Dec 21, 2020-Sep 5, 2021 | 13 | 3,385 | 31 | 3,367 | 81.8 (47.2, 93.7) | >6 momths, all age, all variant |

**Tabel S3 Fully vaccinated BNT162b2 efficacy (VE) for SARS-CoV-2 variant (Delta) infection in elderly aged ≥ 65 years in studies with a retrospective cohort design.**

| No | First author | Country | Duration | Vaccinated | | Unvaccinated | | VE(%) | Note |
| --- | --- | --- | --- | --- | --- | --- | --- | --- | --- |
|  |  |  |  | Observation | cases | Observation | cases |  |  |
| **Short-term VE for preventing Delta infection (2 weeks-6 months after 2nd dose)** | | | | | | | | | |
| 80 | Tartof et al ^6^ | USA | Dec 14, 2020-Aug 8, 2021 | 252,881 | 199 | 1,186,838 | 994 | 77.0 (73.0, 80.0) | 7-203 days, person-years, all age |
| 81 | Goldberget al ^1,4^ | Israel | Jan 16, 2021-Aug 1, 2021 | 589,060 | 1,345 | — | — | 75.0 (58.0, 85.0) | <6 months, 60+ |
| **Short-term VE for preventing severe COVID-19 disease (2 weeks-6 months after 2nd dose)** | | | | | | | | | |
| 80 | Tartof et al ^6^ | USA | Dec 14, 2020-Aug 8, 2021 | 261,945 | 7 | 1,234,999 | 92 | 93.0 (84.0, 96.0) | 7-203 days, person-years, all age |
| 81 | Goldberget al ^1,4^ | Israel | Jan 16, 2021-Aug 1, 2021 | 589,060 | 120 | — | — | 91.0 (85.0, 95.0) | < 6 months, 60+, |
|  | Sheikh et al ^35^ | Scotland | Apr 1, 2021-Aug 16, 2021 | 351 | 24 | 81.4 | 24 | 87.0 (77.0, 93.0) | >14 days, 60+, against death |
| **Long-term VE for preventing Delta infection (>6 months after 2nd dose)** | | | | | | | | | |
| 75 | Nanduri et al ^36^ | USA | Mar 1, 2021-Aug 1, 2021 | 3,248,732 | 1,939 | 953,861 | 1,397 | 52.4 (48.0, 56.4) | ~6 months, nursing home residents |
| 81 | Goldberget al ^1,4^ | Israel | Jan 16, 2021-Aug 1, 2021 | 705,313 | 2,266 | — | — | 57.0 (52.0, 62.0) | ~6 months, 60+ |
| **Long-term VE for preventing severe COVID-19 disease (>6 months after 2nd dose)** | | | | | | | | | |
| 81 | Goldberget al ^1,4^ | Israel | Jan 16, 2021-Aug 1, 2021 | 705,313 | 207 | — | — | 86.0 (82.0, 90.0) | ~6 months, 60+ |

Consolidated Health Economic Evaluation Reporting Standards (CHEERS) Checklist

Items to include when reporting economic evaluations of health interventions

The ISPOR CHEERS Task Force Report, Consolidated Health Economic Evaluation Reporting Standards (CHEERS)—Explanation and Elaboration: A Report of the ISPOR Health Economic Evaluations Publication Guidelines Good Reporting Practices Task Force, provides examples and further discussion of the 24-item CHEERS Checklist and the CHEERS Statement. It may be accessed via the Value in Health or via the ISPOR Health Economic Evaluation Publication Guidelines –CHEERS: Good Reporting Practices webpage:

<http://www.ispor.org/TaskForces/EconomicPubGuidelines.asp>

|  |  | Reporting Item | Page Number |
| --- | --- | --- | --- |
| Title |  |  |  |
|  | [#1](https://www.goodreports.org/reporting-checklists/cheers/info/#1) | Identify the study as an economic evaluation or use more specific terms such as “cost-effectiveness analysis”, and describe the interventions compared. | 1 |
| Abstract |  |  |  |
|  | [#2](https://www.goodreports.org/reporting-checklists/cheers/info/#2) | Provide a structured summary of objectives, perspective, setting, methods (including study design and inputs), results (including base case and uncertainty analyses), and conclusions | 2 |
| Introduction |  |  |  |
| Background and objectives | [#3](https://www.goodreports.org/reporting-checklists/cheers/info/#3) | Provide an explicit statement of the broader context for the study. Present the study question and its relevance for health policy or practice decisions | 3 |
| Methods |  |  |  |
| Target population and subgroups | [#4](https://www.goodreports.org/reporting-checklists/cheers/info/#4) | Describe characteristics of the base case population and subgroups analysed, including why they were chosen. | 3 |
| Setting and location | [#5](https://www.goodreports.org/reporting-checklists/cheers/info/#5) | State relevant aspects of the system(s) in which the decision(s) need(s) to be made. | 3 |
| Study perspective | [#6](https://www.goodreports.org/reporting-checklists/cheers/info/#6) | Describe the perspective of the study and relate this to the costs being evaluated. | 3 |
| Comparators | [#7](https://www.goodreports.org/reporting-checklists/cheers/info/#7) | Describe the interventions or strategies being compared and state why they were chosen. | 4 |
| Time horizon | [#8](https://www.goodreports.org/reporting-checklists/cheers/info/#8) | State the time horizon(s) over which costs and consequences are being evaluated and say why appropriate. | 3-4 |
| Discount rate | [#9](https://www.goodreports.org/reporting-checklists/cheers/info/#9) | Report the choice of discount rate(s) used for costs and outcomes and say why appropriate | 4 |
| Choice of health outcomes | [#10](https://www.goodreports.org/reporting-checklists/cheers/info/#10) | Describe what outcomes were used as the measure(s) of benefit in the evaluation and their relevance for the type of analysis performed | 4 |
| Meaurement of effectiveness | [#11a](https://www.goodreports.org/reporting-checklists/cheers/info/#11a) | Single study-based estimates: Describe fully the design features of the single effectiveness study and why the single study was a sufficient source of clinical effectiveness data | n/a |
| Measurement of effectiveness | [#11b](https://www.goodreports.org/reporting-checklists/cheers/info/#11b) | Synthesis-based estimates: Describe fully the methods used for identification of included studies and synthesis of clinical effectiveness data | 4 |
| Measurement and valuation of preference based outcomes | [#12](https://www.goodreports.org/reporting-checklists/cheers/info/#12) | If applicable, describe the population and methods used to elicit preferences for outcomes. | n/a |
| Estimating resources and costs | [#13a](https://www.goodreports.org/reporting-checklists/cheers/info/#13a) | Single study-based economic evaluation: Describe approaches used to estimate resource use associated with the alternative interventions. Describe primary or secondary research methods for valuing each resource item in terms of its unit cost. Describe any adjustments made to approximate to opportunity costs | n/a |
| Estimating resources and costs | [#13b](https://www.goodreports.org/reporting-checklists/cheers/info/#13b) | Model-based economic evaluation: Describe approaches and data sources used to estimate resource use associated with model health states. Describe primary or secondary research methods for valuing each resource item in terms of its unit cost. Describe any adjustments made to approximate to opportunity costs. | 4 |
| Currency, price date, and conversion | [#14](https://www.goodreports.org/reporting-checklists/cheers/info/#14) | Report the dates of the estimated resource quantities and unit costs. Describe methods for adjusting estimated unit costs to the year of reported costs if necessary. Describe methods for converting costs into a common currency base and the exchange rate. | 4 |
| Choice of model | [#15](https://www.goodreports.org/reporting-checklists/cheers/info/#15) | Describe and give reasons for the specific type of decision analytical model used. Providing a figure to show model structure is strongly recommended. | 3-4 Figure S1 |
| Assumptions | [#16](https://www.goodreports.org/reporting-checklists/cheers/info/#16) | Describe all structural or other assumptions underpinning the decision-analytical model. | 3-4 |
| Analytical methods | [#17](https://www.goodreports.org/reporting-checklists/cheers/info/#17) | Describe all analytical methods supporting the evaluation. This could include methods for dealing with skewed, missing, or censored data; extrapolation methods; methods for pooling data; approaches to validate or make adjustments (such as half cycle corrections) to a model; and methods for handling population heterogeneity and uncertainty. | 4 |
| Results |  |  |  |
| Study parameters | [#18](https://www.goodreports.org/reporting-checklists/cheers/info/#18) | Report the values, ranges, references, and, if used, probability distributions for all parameters. Report reasons or sources for distributions used to represent uncertainty where appropriate. Providing a table to show the input values is strongly recommended. | Table S1 |
| Incremental costs and outcomes | [#19](https://www.goodreports.org/reporting-checklists/cheers/info/#19) | For each intervention, report mean values for the main categories of estimated costs and outcomes of interest, as well as mean differences between the comparator groups. If applicable, report incremental cost-effectiveness ratios. | 5 |
| Characterising uncertainty | [#20a](https://www.goodreports.org/reporting-checklists/cheers/info/#20a) | Single study-based economic evaluation: Describe the effects of sampling uncertainty for the estimated incremental cost and incremental effectiveness parameters, together with the impact of methodological assumptions (such as discount rate, study perspective). | n/a |
| Characterising uncertainty | [#20b](https://www.goodreports.org/reporting-checklists/cheers/info/#20b) | Model-based economic evaluation: Describe the effects on the results of uncertainty for all input parameters, and uncertainty related to the structure of the model and assumptions. | 5 Figure 1 |
| Characterising heterogeneity | [#21](https://www.goodreports.org/reporting-checklists/cheers/info/#21) | If applicable, report differences in costs, outcomes, or cost effectiveness that can be explained by variations between subgroups of patients with different baseline characteristics or other observed variability in effects that are not reducible by more information. | 5 |
| Discussion |  |  |  |
| Study findings, limitations, generalisability, and current knowledge | [#22](https://www.goodreports.org/reporting-checklists/cheers/info/#22) | Summarise key study findings and describe how they support the conclusions reached. Discuss limitations and the generalisability of the findings and how the findings fit with current knowledge. | 6-7 |
| Other |  |  |  |
| Source of funding | [#23](https://www.goodreports.org/reporting-checklists/cheers/info/#23) | Describe how the study was funded and the role of the funder in the identification, design, conduct, and reporting of the analysis. Describe other non-monetary sources of support | 8 |
| Conflict of interest | [#24](https://www.goodreports.org/reporting-checklists/cheers/info/#24) | Describe any potential for conflict of interest of study contributors in accordance with journal policy. In the absence of a journal policy, we recommend authors comply with International Committee of Medical Journal Editors recommendations | 8 |

For consistency, the CHEERS Statement checklist format is based on the format of the CONSORT statement checklist

The ISPOR CHEERS Task Force Report provides examples and further discussion of the 24-item CHEERS Checklist and the CHEERS Statement. It may be accessed via the Value in Health link or via the ISPOR Health Economic Evaluation Publication Guidelines – CHEERS: Good Reporting Practices

webpage: http://www.ispor.org/TaskForces/EconomicPubGuidelines.asp

The citation for the CHEERS Task Force Report is: Husereau D, Drummond M, Petrou S, et al. Consolidated health economic evaluation reporting standards (CHEERS)—Explanation and elaboration: A report of the ISPOR health economic evaluations publication guidelines good reporting practices task force. Value Health 2013; 16:231–50.
